# Validity, reliability, and user perspectives of the newly developed joint angle measurement system with inertial measurement unit sensors

**DOI:** 10.1101/2025.04.24.25326137

**Authors:** Taiki Yoshida, Shintaro Uehara, Asuka Hirano, Shota Itoh, Yohei Otaka

## Abstract

**Purpose:** We aimed to evaluate the applicability of the newly developed joint angle measurement system consisting of six-axis inertial measurement unit sensors and tablet-based application that measure and store angular velocity and acceleration data for estimating joint angles.

**Materials and Methods:** The tablet-based application was used to calculate the orientation angles from angular velocity and acceleration data measured using a single sensor. The relative angles were calculated using the data from multiple sensors. In experiment 1, the validity and reliability of calculated angles was examined using a test device. In experiment 2, the angles of five joints were calculated in four healthy participants using attached inertial measurement unit sensors; the angles were compared with universal goniometer-measured values. In experiment 3, usability and satisfaction were evaluated with the System Usability Scale (SUS) and a Quebec User Evaluation of Satisfaction with Assistive Technology (QUEST)-like scale.

**Results:** In experiment 1, the mean difference of the time-series data between those obtained by the developed system and test device was ˂0.2° for all axes. In experiment 2, the mean difference of the integrated data was 0.2°. The mean difference for all joints was ˂5°, indicating that the measurement system is comparable to the universal goniometer. In experiment 3, the median SUS and QUEST-like scale scores were 81 and 4.0, respectively, indicating high usability and satisfaction.

**Conclusion:** The newly developed joint angle measurement system has high accuracy in measuring angles and sufficient validity in application to human joint angles, with high usability and user satisfaction.

## Introduction

Identifying the differences between normal and pathological movements to understand the characteristics of impairments is essential for planning rehabilitation exercise and evaluating its effectiveness. Therefore, rehabilitation professionals often use kinematic assessments such as joint angle measurement. Three-dimensional (3D) motion analysis systems using optical markers are commonly used for kinematic measurements, and they have shown good inter-rater variability and allow for the tracking of joint angle changes during active movements (1). However, this method requires multiple infrared cameras, thus limiting its use to the laboratory (2,3). Consequently, the measurement of patients’ activities in real-life and clinical settings, such as hospital wards, homes, and outdoor environments, appears impractical. Therefore, there is a need to develop simpler measurement methods as alternatives to 3D motion analysis systems for kinematic measurements in real-life settings.

In recent years, inertial measurement unit (IMU) sensors have gained attention in the field of rehabilitation as a convenient method for measuring joint angles (4–6), potentially providing a solution to the limitations of 3D motion analysis systems. IMU sensors typically comprise accelerometers, gyroscopes, etc. to estimate posture and joint angles. Previous studies have reported the reliability and validity of IMU sensors for the measurement of the joint angles in the neck (7), shoulder (8–11), elbow (10–13), wrist (10,11), hip (14,15), knee (16,17), and trunk (18,19). In addition, similar to the 3D motion analysis system using optical markers, IMU-based measurements allow the continuous tracking of joint movements during active motion (20). Therefore, IMU sensors can become a better alternative method for measuring human joint angles in rehabilitation settings. However, most studies on the applicability of IMU sensors in rehabilitation settings were performed at the laboratory level, and there is limited research on their feasibility for daily clinical use in real-world settings. This may be due to the lack of user-friendly applications for analysis in rehabilitation settings and/or the limited availability of devices that can measure the upper and lower limb joint angles using a single measurement system (21). To address these issues, we developed a new joint angle measurement system consisting of IMU sensors and tablet-based user-friendly application that enables easy measurement, data storage, and analysis of joint angles in the upper limbs, lower limbs, and trunk.

In the present study, we aimed to evaluate the clinical feasibility of the newly developed joint angle measurement system by investigating the validity and reliability of the calculated angles, usability of the system, and user satisfaction. Specifically, in experiment 1, we investigated whether the developed system could accurately measure orientation angles. In experiment 2, we calculated joint angles in static positions for five joints in the upper, lower limbs, and trunk using the IMU sensors attached to the human body to examine the bias in the angles from those measured by the universal goniometer. In experiment 3, we evaluated the usability and user satisfaction of the newly developed measurement system using questionnaires in rehabilitation professionals.

## Materials and Methods

### Newly developed joint angle measurement system

We developed a joint angle measurement system consisting of six-axis IMU sensors and an angle-calculating application (Figure 1). First, the IMU sensors acquire angular velocity and acceleration data in the local coordinate system relative to the sensor body. Thereafter, the data are fused using a specific algorithm to calculate the orientation angles (roll, pitch, and yaw) of the sensor. Subsequently, the orientation angles are transformed into the world coordinate system, and the relative rotational angles between multiple sensors are computed to derive the joint angles. The calculated angle data are transmitted in real-time via Bluetooth low energy to a connected tablet device (iPad 3rd Gen. Apple Store, Cupertino, Ca USA) with a sampling frequency of 20 Hz, where the analyzed results are displayed on a developed application.

**Figure 1.**
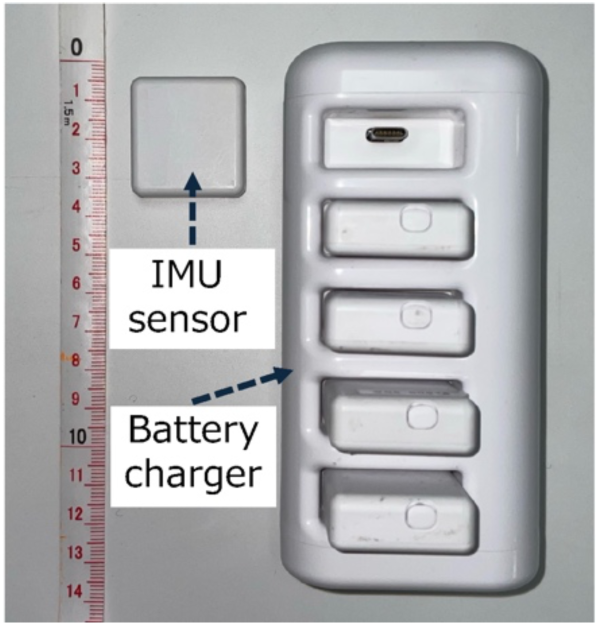
Overview of the developed angle measurement system. The system consists of IMU sensors, a battery charger, and a tablet-based application. IMU: inertial measurement unit

### Experiment 1: Accuracy of the calculated angles

To verify the accuracy of the calculated angles obtained using the developed measurement system, we used a test device that repetitively moved at specified angles (Figure 2). The device performs repetitive rotating movements from a neutral position of 0°, reaching up to ±42.5° to each side. As it approaches the maximum (+42.5°) or minimum angle (−42.5°), the rotational speed gradually decreases. One complete cycle of movement (from +42.5° to −42.5° and back) takes approximately 8 s. The IMU sensor was secured to ensure the alignment of its rotational axis aligned with the roll, pitch, and yaw axes, and each axis underwent 10 cycles of back-and-forth rotation.

**Figure 2.**
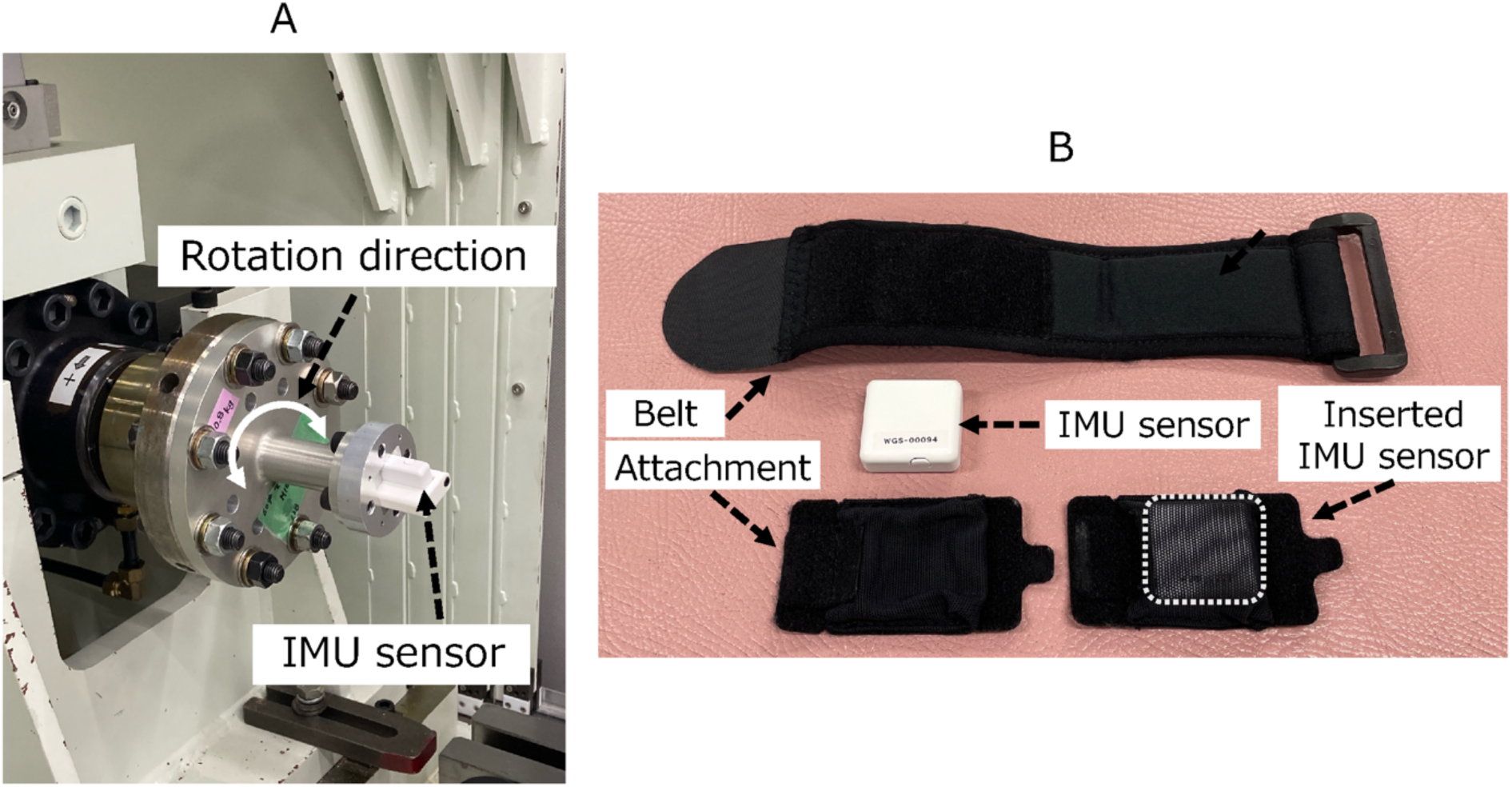
(A) Test device used in the Experiment 1. The test device repeatedly rotates back and forth, with a baseline of 0°, reaching 42.5° on each side. **(B)** The specialized attachment used in the study. The IMU sensor was inserted in the attachment, which was secured to the belt worn on the body with the Velcro fastener. IMU: inertial measurement unit

First, to investigate the validity of the calculated angles, we analyzed the bias in the angles obtained over time from the test device and measurement system during the 10 cycles of rotation using Bland–Altman analysis. Next, to confirm the reliability of the calculated angles, we focused on the angles obtained at the transition points of the rotating directions (i.e., the maximum and minimum angles) of the test device. Two sets of 10 cycles of rotation were measured (first and second sets) and the differences in the 10 maximum and minimum angles between the first and second sets were evaluated using Bland–Altman analysis. We calculated angle differences, standard deviation (SD), and 95% confidence interval (95% CI) of the bias, as well as the Upper Limits of Agreement (ULoA) and its 95% CI and the Lower Limits of Agreement (LLoA) and its 95% CI. The presence of fixed bias was confirmed when the 95% CI of the differences between the two measurement methods or two sessions did not include zero. Proportional bias was tested using a linear regression analysis and confirmed when the regression coefficient was not equal to zero (p < 0.05). R (version 4.4.2) was used for data analysis.

### Experiment 2: Accuracy of the system for human joint angle measurement

Experiment 2 was conducted with the approval of the Ethics Review Committee of Fujita Health University (approval number: HM21-029) and in accordance with the Declaration of Helsinki of 1964, as revised in 2013. Before conducting the measurements, the researchers provided the participants with an oral explanation and written materials detailing the study, and written consent was obtained from each participant prior to the experiments.

Experiment 2 aimed to verify the accuracy of the calculated angles when measuring the human joint angle at several static positions by comparing them with those measured using the universal goniometer, a commonly used joint angle measurement device in clinical settings. Four healthy volunteers (two females) with a mean age (standard deviation [SD]) of 29.0 (4.0) years participated in the experiment. The IMU sensors were attached to the following locations with a specialized attachment (Figure 2): the posterior part of the mid-upper arm, the dorsal part of the distal forearm, the back at the level of the first thoracic vertebra (spinous process) and at the level between the spinous processes of the fourth and fifth lumbar vertebrae (intersection of the Jacoby’s line and the spine), the anterior part of the mid-thigh, and the anterior part of the distal lower leg.

Participants held the arbitrary position at 10 different angles for each of the five joint movements: shoulder flexion, elbow flexion, hip flexion, knee flexion, and trunk flexion. The angle of the five joint movements were calculated using the following sensor combinations: shoulder flexion, sensors on the posterior mid-upper arm and the back at the first thoracic vertebra; elbow flexion, sensors on the posterior mid-upper arm and dorsal distal forearm; hip flexion, sensors between the fourth and fifth lumbar vertebrae and anterior mid-thigh; knee flexion, sensors on the anterior mid-thigh and anterior distal lower leg; and trunk flexion, sensors on the back at the level of the first thoracic vertebra and at the level between the spinous processes of the fourth and fifth lumbar vertebrae. To compare the calculated angles of the measurement system with those measured using the universal goniometer, two raters—an occupational therapist with 15 years of clinical experience and a physical therapist with 21 years of clinical experience—measured the joint angles with the universal goniometer while simultaneously measuring the static joint angles with the measurement system. Both raters checked the angle measured using the universal goniometer and reached a consensus on each measurement value (1° increment).

The analysis was first performed by integrating all measured ten angles of the five joints from all four participants; afterward, the bias of angles between the measurement system and universal goniometer were examined using Bland–Altman analysis. Subsequently, the same analysis was performed separately for the angles of each joint. We calculated the difference of angles, SD, 95% CI of the bias, ULoA and its 95% CI, and LLoA and its 95% CI. The presence of a fixed bias was confirmed when the 95% CI of the differences between the two measurement methods did not include zero. Proportional bias was tested using a linear regression analysis and confirmed when the regression coefficient was not equal to zero (p < 0.05). R (version 4.4.2) was used for the analysis.

### Experiment 3: Evaluation of usability and user satisfaction

Experiment 3 was conducted following the same procedure as that of Experiment 2, with the approval of the Ethical Review Committee and consent of the participants. Ten rehabilitation professionals from the Department of Rehabilitation at Fujita Health University Hospital participated in this experiment: seven physical therapists (one female; mean [SD] years of clinical experience, 7.6 [3.0]) and three occupational therapists (one female; mean [SD] years of clinical experience, 7.7 [1.2]). After a detailed explanation of how to use the developed measurement system, participants were asked to perform a set of standardized tasks using the developed system, including sensor attachment, operation of the tablet application, and data retrieval. They were allowed sufficient time (approximately 60 min) to familiarize themselves with the developed system through hands-on experience. Following this session, participants completed the System Usability Scale (SUS) (22) and a modified version of the Quebec User Evaluation of Satisfaction with Assistive Technology (QUEST) (23) questionnaires to evaluate the usability of and satisfaction with the system. The SUS is a reliable measure for assessing the usability of various products and services. It consists of 10 items scored on a scale of 0 to 100, with participants rating each item on a five-point Likert scale, ranging from “strongly agree” to “strongly disagree.” Higher scores indicate greater usability of the system. An SUS score below 50 indicates that users rated the system as “OK,” whereas scores of 70 or higher are considered “good” and scores above 85 are deemed “excellent.” The QUEST is a reliable measure for evaluating user satisfaction with assistive technology. It consists of items regarding assistive devices and services. In the present study, we only used the items regarding assistive devices to better suit the evaluation of the developed system: size (height, length, and width), weight, ease of adjustment (installation and adjustment of components), safety, durability, ease of use, comfort, and effectiveness. Each aspect was rated on a five-point Likert scale, comprising “1: Not satisfied at all”, “2: Not very satisfied”, “3: more or less satisfied”, “4: Quite satisfied”, and “5: very satisfied.” The mean score across all items was calculated as the overall satisfaction score for the system, and this score was interpreted with higher scores reflecting higher levels of satisfaction.

## Results

### Experiment 1

The Bland–Altman plot of time-series data obtained using the measurement system and test device (i.e., validity relative to the angles obtained from the test device) revealed a statistically significant fixed bias for the Roll and Yaw axes (p < 0.001) and proportional bias in the Pitch and Roll axes (p < 0.001). However, the mean differences were ˂0.2° for all axes, and the ULoA and LLoA were ˂2° (Figure 3, Table 1). In addition, the Bland–Altman plot of the maximum and minimum angles between the first and second sets of 10 cycles of rotating movements obtained using the measurement system (i.e., reliability of the calculated angles) revealed a statistically significant fixed bias in the minimum Roll and Pitch angles (p = 0.039 and p = 0.028, respectively) and a proportional bias in the maximum Pitch and minimum Yaw angles (p = 0.009 and p <0.001, respectively). The mean differences were ˂0.1° for all axes, with their ULoA and LLoA ˂1° (Table 2). These results suggest that, despite the presence of statistically significant fixed and proportional biases in some axes, the magnitude of these biases was minimal from a clinical perspective. Therefore, the calculated angles can be considered valid and reliable.

**Figure 3.**
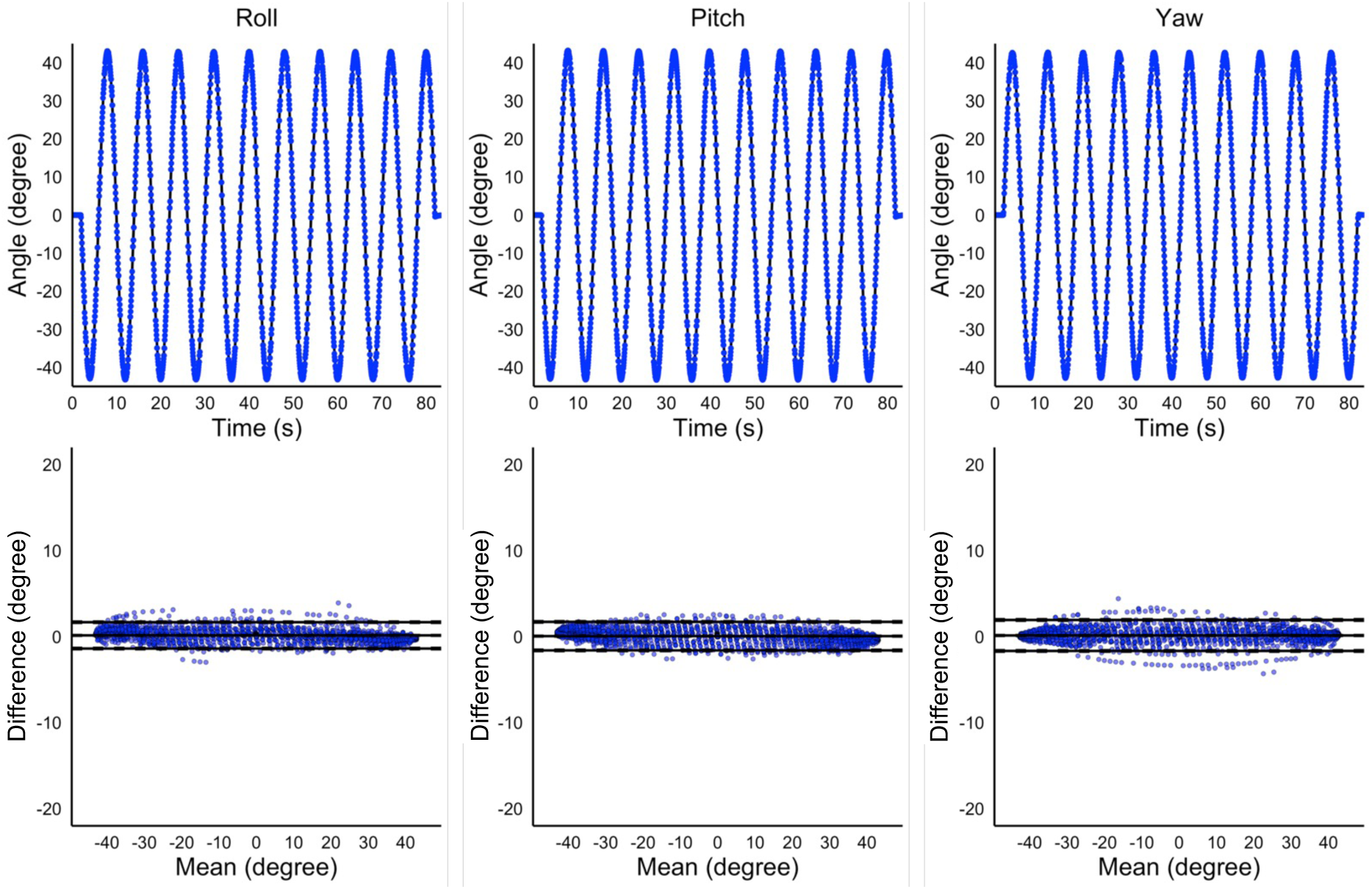
Biases of the measured joint angles between the developed system and test device. The top three panels represent the time-series of angles of the test device (solid lines) and output angles measured using the developed system (dotted lines). The X and Y axes show the time and the measured angle, respectively. The bottom three panels represent the biases in the measured angle between the two measurement methods using the Bland–Altman plots. Each plot represents the angle of the test device minus the angle calculated by the developed system. X and Y axes show the mean value of the two measurement methods and the bias between them, respectively. The middle solid line represents the bias; the top and bottom solid lines show the Limits of Agreement. The dotted lines show the 95% confidence intervals of the bias and the Limits of Agreement.

**Table 1.** Bias of the time-series angles between those calculated using the developed system and test device.

|  | Roll | Pitch | Yaw |
| --- | --- | --- | --- |
| Fixed bias |  |  |  |
| Mean difference | 0.14 | 0.05 | 0.13 |
| SD | 0.84 | 0.78 | 0.91 |
| 95% CI | 0.10 – 0.18 | 0.01 – 0.09 | 0.08 – 0.17 |
| Upper LoA | 1.67 | 1.71 | 1.93 |
| 95% CI | 1.59 – 1.75 | 1.62 – 1.80 | 1.83 – 2.03 |
| Lower LoA | -1.38 | -1.60 | -1.67 |
| 95% CI | -1.47 – -1.30 | -1.69 – -1.51 | -1.76 – -1.57 |
| p-value | <0.001 | 0.007 | <0.001 |
| Proportional bias |  |  |  |
| r | -0.008 | -0.009 | <0.001 |
| p-value | <0.001 | <0.001 | 0.366 |

SD: Standard deviation; LoA: Limits of Agreement; CI: confidence interval

**Table 2.** Bias of the maximum and minimum angle between the first and second sets of 10 cycles of rotational motion calculated using the measurement system.

|  | Maximum angles |  |  | Minimum angles |  |  |
| --- | --- | --- | --- | --- | --- | --- |
|  | Roll | Pitch | Yaw | Roll | Pitch | Yaw |
| Fixed bias |  |  |  |  |  |  |
| Mean difference | 0.01 | 0.08 | 0.01 | 0.01 | 0.01 | -0.06 |
| SD | 0.02 | 0.18 | 0.04 | 0.01 | 0.01 | 0.29 |
| 95% CI | 0.00 – 0.02 | -0.05 – 0.21 | -0.01 – 0.04 | 0.00 – 0.02 | 0.00 – 0.03 | -0.27 – 0.14 |
| Upper LoA | 0.06 | 0.43 | 0.09 | 0.05 | 0.05 | 0.50 |
| 95% CI | 0.02 – 0.10 | 0.15 – 0.72 | 0.03 – 0.16 | 0.02 – 0.08 | 0.02 – 0.08 | 0.04 – 0.96 |
| Lower LoA | -0.03 | -0.27 | -0.06 | -0.02 | -0.02 | -0.64 |
| 95% CI | -0.07 – 0.00 | -0.56 – 0.01 | -0.12 – 0.00 | -0.05–0.00 | -0.05 – 0.00 | -1.10 – -0.18 |
| p-value | 0.195 | 0.195 | 0.190 | 0.039 | 0.028 | 0.4757 |
| Proportional bias |  |  |  |  |  |  |
| r | -0.030 | -1.138 | -0.672 | 0.003 | -0.060 | -1.824 |
| p-value | 0.850 | 0.009 | 0.219 | 0.970 | 0.485 | <0.001 |

SD: Standard deviation; LoA: Limits of Agreement; CI: Confidence interval

### Experiment 2

In the Bland-Altman plot comparing the developed system with the universal goniometer using the integrated dataset comprising ten angles of five joints from four participants, a statistically significant proportional bias was observed (p = 0.016); however, no fixed bias was confirmed. The mean differences were 0.2°, with a ULoA of 7.8° and an LLoA of −7.3°. Analysis of each individual joint revealed a statistically significant fixed bias in all joints (p < 0.05), and a proportional bias was observed in the shoulder, hip, and knee joints (p < 0.05). However, even the largest mean difference was 3.4° in the trunk, and the largest ULoA and LLoA were 3.9° and −8.8°, respectively, in the knee joint, indicating minimal differences when considering potential effects in the clinical setting. These results suggest that, despite the presence of some fixed or proportional biases, the joint angles calculated using the developed measurement system have acceptable validity relative to those obtained using the universal goniometer (Figure 4, Table 3).

**Figure 4.**
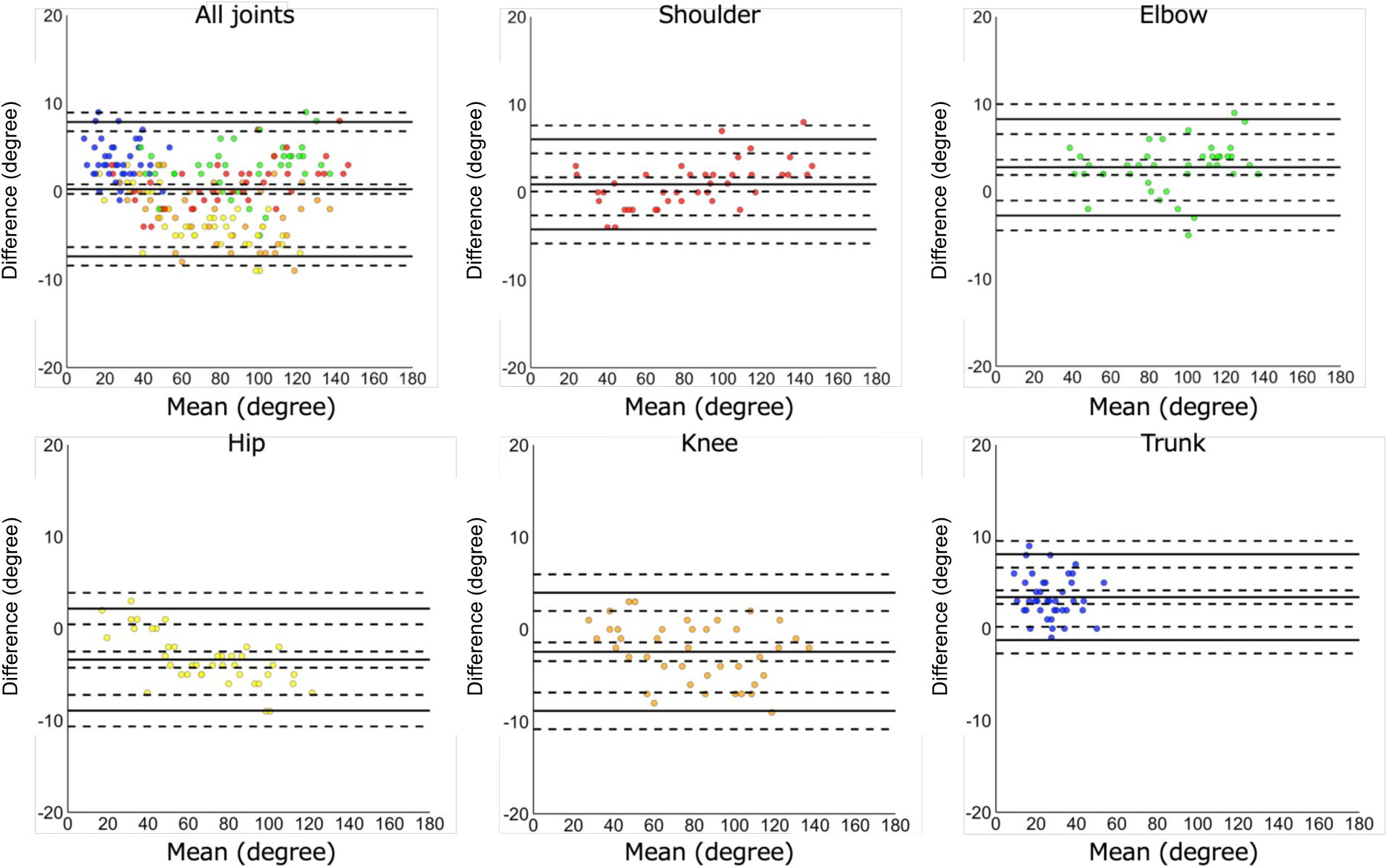
Biases of the measured joint angles between the developed system and universal goniometer. Each plot represents the angle measured by the universal goniometer minus the angle calculated by the developed system. The top left panel represents the biases in the measured angles between the two methods using the Bland–Altman plots, with all joint movement data integrated. The other five panels represent the biases for each joint: shoulder (top center), elbow (top right), hip (bottom left), knee (bottom center), and trunk (bottom right). X and Y axes show the mean value of the two measurement methods and the bias between them, respectively. The middle solid line represents the bias; the top and bottom solid lines show the Limits of Agreement. The dotted lines show the 95% confidence intervals of the bias and the Limits of Agreement.

**Table 3.** Bias of the joint angles between those measured using the developed system and universal goniometer.

|  | All joints | Shoulder | Elbow | Hip | Knee | Trunk |
| --- | --- | --- | --- | --- | --- | --- |
| Fixed bias |  |  |  |  |  |  |
| Mean difference | 0.2 | 0.9 | 2.7 | -3.3 | -2.4 | 3.4 |
| SD | 3.8 | 2.6 | 2.8 | 2.8 | 3.2 | 3.4 |
| 95% CI | -0.2 – 0.8 | 0.0 – 1.7 | 1.8 – 3.6 | -4.2 – -2.4 | -3.4 – -1.3 | 2.6 – 4.1 |
| Upper LoA | 7.8 | 6.0 | 8.2 | 2.1 | 3.9 | 8.1 |
| 95% CI | 6.7 – 9.0 | 4.1 – 7.8 | 6.3 – 10.2 | 0.1 – 4.1 | 1.6 – 6.2 | 6.4 – 9.7 |
| Lower LoA | -7.3 | -4.2 | -2.7 | -8.9 | -8.8 | -1.2 |
| 95% CI | -8.5 – -6.1 | -6.0 – -2.8 | -4.7 – -0.7 | -10.9 – -6.9 | -11.1 – -6.5 | -2.9 – 0.4 |
| p-value | 0.346 | 0.035 | <0.001 | <0.001 | <0.001 | <0.001 |
| Proportional bias |  |  |  |  |  |  |
| r | -0.018 | 0.039 | 0.022 | -0.072 | -0.034 | -0.030 |
| p-value | 0.016 | <0.001 | 0.160 | <0.001 | 0.046 | 0.401 |

SD: Standard deviation; LoA: Limits of Agreement; CI: Confidence interval

### Experiment 3

Although there were some differences between individuals, the median SUS score was 81.3 (interquartile range [IQR]: 60.0–95.0), demonstrating the usability of the system as “good.” The median score for the QUEST-like scale was 4.0 (IQR: 3.8–4.3), indicating that the participants were “quite satisfied” with the system. Further analysis of the individual items revealed that the median score for all items was above “3.0: more or less satisfied.” These results on user perspectives demonstrate no significant defects in the system (Table 4).

**Table 4.** Scores of usability and user satisfaction of the developed system.

| Participant | 1 | 2 | 3 | 4 | 5 | 6 | 7 | 8 | 9 | 10 | Median |
| --- | --- | --- | --- | --- | --- | --- | --- | --- | --- | --- | --- |
| SUS | 82.5 | 95.0 | 75.0 | 95.0 | 72.5 | 80.0 | 85.0 | 67.5 | 60.0 | 95.0 | 81.3 |
| QUEST-like | 4.4 | 4.3 | 3.9 | 4.6 | 3.9 | 4.1 | 3.8 | 3.6 | 3.4 | 4.4 | 4.0 |
| mean of sub-items |  |  |  |  |  |  |  |  |  |  |  |
| Size | 5 | 4 | 4 | 5 | 5 | 4 | 5 | 4 | 4 | 5 | 4.5 |
| Weight | 5 | 4 | 5 | 5 | 5 | 5 | 5 | 4 | 5 | 5 | 5.0 |
| Ease of adjustment | 4 | 4 | 4 | 5 | 3 | 4 | 4 | 4 | 2 | 4 | 4.0 |
| Safety | 5 | 5 | 4 | 5 | 3 | 5 | 3 | 4 | 4 | 4 | 4.0 |
| Durability | 4 | 4 | 2 | 4 | 3 | 3 | 3 | 3 | 4 | 2 | 3.0 |
| Ease of use | 4 | 5 | 4 | 4 | 5 | 4 | 4 | 3 | 3 | 5 | 4.0 |
| Comfort | 4 | 4 | 4 | 5 | 3 | 4 | 3 | 3 | 2 | 5 | 4.0 |
| Effectiveness | 4 | 4 | 4 | 4 | 4 | 4 | 3 | 4 | 3 | 5 | 4.0 |

SUS: System Usability Scale; QUEST-like: Quebec User Evaluation of Satisfaction with Assistive Technology-like questionnaire

## Discussion

In the present study, for evaluating the clinical feasibility of the newly developed joint angle measurement system consisting of the IMU sensors and tablet-based application, we investigated the validity and reliability of the calculated values, usability, and user satisfaction. The results showed that the developed system can accurately capture angular changes, demonstrating good usability and user satisfaction. These findings indicate that the newly developed measurement system has a high potential for clinical application in measuring joint range of motion in the upper limbs, lower limbs, and trunk.

In experiment 1, the validity and reliability of the angle measurements obtained from the developed measurement system was assessed using the test device. The results confirmed that the developed measurement system is capable of measuring highly variable and reliable temporal angle data. In experiment 2, we investigated the bias in the measured angles of the measurement system and universal goniometer to confirm the validity of the measurement system in measuring human joint angles. The results showed that the mean difference of the integrated data obtained using the measurement system and universal goniometer was ˂1°. Moreover, focusing on individual joints, the bias for all joints was ˂5°. These results were comparable to those of previous studies comparing joint angles measured using IMU sensors and universal goniometer (8,9,13,15), indicating that the level of bias was within an acceptable range for clinical applications.

However, the mean difference in the measured angles found in experiment 2 was greater than that observed in experiment 1, where the calculated angles were compared with those of the test device. The mean difference in joint angles measured using the developed system and universal goniometer is likely due to biases in the definitions of reference axes for each measure. The axes for the universal goniometer are usually a bone and a reference line in the external environment. In contrast, the axes for the IMU sensor are the body of the device itself. For example, for shoulder flexion, the universal goniometer defines a vertical line to the floor passing through the acromion as the reference axis and the humerus as the other axis (24). In the developed measurement system, the angle formed between the sensor attached to the back and the sensor attached to the upper arm is defined as shoulder flexion. This mean difference may be a major contributor to the discrepancies in the measured angles, specifically in the following scenarios. The first scenario is when the muscle mass or amount of adipose tissue at the attachment site changes due to the expansion and contraction of these tissues along with changes in joint angles. This assumption can be supported by the result of the present study showing that angle bias was larger for lower limb joints and trunk, which have more muscle mass and adipose tissue amount, leading to a greater likelihood of the sensor to shift compared to those for upper limb joints. In addition, this result indicates that caution should be applied to areas with greater muscle mass and adipose tissue amount and careful consideration is needed ensure a secure attachment when measuring joint angles using the developed system. The second scenario is when the IMU sensor attached to the body moves due to insecure attachment, resulting in misalignment of the sensor’s position. In this study, special attachments were used to secure the sensors to the body to reduce the risk of misalignment (Figure 2). However, it is still possible that slight sensor misalignment could not be entirely prevented. It has been also demonstrated that, when using optical markers attached to the body surface for human motion analysis, misalignment of the markers cannot be eliminated, leaving measuring errors/noise to be removed (25). Therefore, there is need for a better protocol to secure the IMU sensor to the body.

In the present study, in addition to evaluating the validity and reliability of the developed system for angle measurement, we evaluated the usability and user satisfaction of the system in experiment 3 from the perspective of practical clinical application. User experience, a comprehensive concept that includes usability, is essential for promoting the clinical adoption of medical devices and enhancing their effectiveness and safety (26,27). In this regard, the present study confirmed that the newly developed measurement system had high usability and user satisfaction. Therefore, it can be concluded that the developed system has sufficient potential for clinical application.

This study had several limitations. Regarding the validity of the developed system compared to the universal goniometer, the number of participants and the variety of joints and movements assessed were limited. Individual differences in body composition—such as the amount of muscle mass and adipose tissue—may have caused a bias in the calculated joint angles. Future studies should include participants with diverse body shapes and assess various joints and movements. Moreover, the present study focused on the validity of joint angles in static positions. To apply the developed system in real-life and clinical settings for motion analysis, future investigations will be needed to validate the joint angles during active movements.

## Conclusions

The present study evaluated the validity, reliability, usability, and user satisfaction of the newly developed joint angle measurement system consisting of IMU sensors and tablet-based application, confirming its potential for clinical application. The use of the developed system offers the possibility of extending motion analysis from the laboratory settings to more natural and practical environments, such as hospital wards, homes, and outdoor environments. However, further clinical validations are required to establish the applicability of the system.

## Acknowledgments

The authors thank Mr. Makoto Kobayashi and Mr. Haruki Inoue at TOYOTA Motor Corporation for providing technical support for the development of the device used.

## Data availability statement

The original contributions presented in the study are included in the article/Supplemental data, further inquiries can be directed to the corresponding author.

## References

1. Moeslund TB, Granum E. A survey of computer vision-based human motion capture. Comput Vis Image Underst. 2001 Mar;81(3):231–68. doi: 10.1006/cviu.2000.0897

2. Wong WY, Wong MS, Lo KH. Clinical applications of sensors for human posture and movement analysis: a review. Prosthet Orthot Int. 2007 Mar;31(1):62–75. doi: 10.1080/03093640600983949.

3. Gu C, Lin W, He X, Zhang L, Zhang M. IMU-based motion capture system for rehabilitation applications: A systematic review. Biomimetic Intelligence and Robotics. 2023 Jun 1;3(2):100097. doi: 10.1016/j.birob.2023.100097

4. Cuesta-Vargas AI, Galán-Mercant A, Williams JM. The use of inertial sensors system for human motion analysis. Phys Ther Rev. 2010 Dec;15(6):462–73. doi: 10.1179/1743288X11Y.0000000006

5. Lefeber N, Degelaen M, Truyers C, Safin I, Beckwee D. Validity and reproducibility of inertial Physilog sensors for spatiotemporal gait analysis in patients with stroke. IEEE Trans Neural Syst Rehabil Eng. 2019 Sep;27(9):1865–74. doi: 10.1109/TNSRE.2019.2930751

6. Patel G, Mullerpatan R, Agarwal B, Shetty T, Ojha R, Shaikh-Mohammed J, et al. Validation of wearable inertial sensor-based gait analysis system for measurement of spatiotemporal parameters and lower extremity joint kinematics in sagittal plane. Proc Inst Mech Eng H. 2022 May;236(5):686–96. doi: 10.1177/09544119211072971

7. English DJ, Weerakkody N, Zacharias A, Green RA, Hocking C, Bini RR. The validity of a single inertial sensor to assess cervical active range of motion. J Biomech. 2023 Oct;159(111781):111781. doi: 10.1016/j.jbiomech.2023.111781

8. Kaszyński J, Baka C, Białecka M, Lubiatowski P. Shoulder range of motion measurement using inertial measurement unit-concurrent validity and reliability. Sensors (Basel) [Internet]. 2023 Aug 29 [cited 2024 Sep 5];23(17). Available from: https://pubmed.ncbi.nlm.nih.gov/37687955/

9. Yoon T-L. Validity and reliability of an inertial measurement unit-based 3D angular measurement of shoulder joint motion. J Korean Phys Ther. 2017 Jun 30;29(3):145–51. doi: 10.18857/jkpt.2017.29.3.145

10. Bouvier B, Duprey S, Claudon L, Dumas R, Savescu A. Upper limb kinematics using inertial and magnetic sensors: Comparison of sensor-to-segment calibrations. Sensors (Basel). 2015 Jul 31;15(8):18813–33. doi: 10.3390/s150818813

11. Pérez R, Costa Ú, Torrent M, Solana J, Opisso E, Cáceres C, et al. Upper limb portable motion analysis system based on inertial technology for neurorehabilitation purposes. Sensors (Basel). 2010 Dec 2;10(12):10733–51. doi: 10.3390/s101210733

12. Santospagnuolo A, Bruno AA, Pagnoni A, Martello F, Santoboni F, Perroni F, et al. Validity and reliability of the GYKO inertial sensor system for the assessment of the elbow range of motion. J Sports Med Phys Fitness. 2019 Sep;59(9):1466–71. doi: 10.23736/S0022-4707.19.09331-9

13. Costa V, Ramírez Ó, Otero A, Muñoz-García D, Uribarri S, Raya R. Validity and reliability of inertial sensors for elbow and wrist range of motion assessment. PeerJ. 2020 Aug 11;8(e9687):e9687. doi: 10.7717/peerj.9687

14. Bakhshi S, Mahoor MH, Davidson BS. Development of a body joint angle measurement system using IMU sensors. Annu Int Conf IEEE Eng Med Biol Soc. 2011;2011:6923–6. doi: 10.1109/IEMBS.2011.6091743

15. Stołowski Ł, Niedziela M, Lubiatowski B, Lubiatowski P, Piontek T. Validity and reliability of inertial measurement units in active range of motion assessment in the hip joint. Sensors (Basel) [Internet]. 2023 Oct 28;23(21). Available from: https://pubmed.ncbi.nlm.nih.gov/37960493/

16. Jaysrichai T, Suputtitada A, Khovidhungij W. Mobile sensor application for kinematic detection of the knees. Ann Rehabil Med. 2015 Aug;39(4):599–608. doi: 10.5535/arm.2015.39.4.599

17. Fennema MC, Bloomfield RA, Lanting BA, Birmingham TB, Teeter MG. Repeatability of measuring knee flexion angles with wearable inertial sensors. Knee. 2019 Jan;26(1):97–105. doi: 10.1016/j.knee.2018.11.002

18. Ali F, Hogen CA, Miller EJ, Kaufman KR. Validation of pelvis and trunk range of motion as assessed using inertial measurement units. Bioengineering (Basel). 2024 Jun 28;11(7):659. doi: 10.3390/bioengineering11070659

19. Bauer CM, Rast FM, Ernst MJ, Kool J, Oetiker S, Rissanen SM, et al. Concurrent validity and reliability of a novel wireless inertial measurement system to assess trunk movement. J Electromyogr Kinesiol. 2015 Oct;25(5):782–90. doi: 10.1016/j.jelekin.2015.06.001

20. García-de-Villa S, Casillas-Pérez D, Jiménez-Martín A, García-Domínguez JJ. Inertial sensors for human motion analysis: A comprehensive review. IEEE Trans Instrum Meas. 2023;72:1–39. doi: 10.48550/arXiv.2401.12919

21. Lopez-Nava IH, Munoz-Melendez A. Wearable inertial sensors for human motion analysis: A review. IEEE Sens J. 2016 Nov;16(22):7821–34. doi: 10.1109/jsen.2016.2609392

22. Bangor A, Kortum P, Miller J. Determining what individual SUS scores mean: Adding an adjective rating scale [Internet]. 2009 [cited 2023 Dec 3]. Available from: https://uxpajournal.org/wp-content/uploads/sites/7/pdf/JUS_Bangor_May2009.pdf

23. Demers L, Weiss-Lambrou R, Ska B. The Quebec User Evaluation of Satisfaction with Assistive Technology (QUEST 2.0): An overview and recent progress. Technol Disabil. 2002 Sep 29;14(3):101–5. doi: 10.13072/midss.298

24. Norkin CC, White DJ. Measurement Of Joint Motion: A Guide To Goniometry. F.A. Davis; 2016.

25. Gao B, Zheng NN. Investigation of soft tissue movement during level walking: translations and rotations of skin markers. J Biomech. 2008 Nov 14;41(15):3189–95. doi: 10.1016/j.jbiomech.2008.08.028

26. Bitkina OV, Kim HK, Park J. Usability and user experience of medical devices: An overview of the current state, analysis methodologies, and future challenges. Int J Ind Ergon. 2020 Mar 1;76(102932):102932. doi: 10.1016/j.ergon.2020.102932

27. Kim T. Factors influencing usability of rehabilitation robotic devices for lower limbs. Sustainability. 2020 Jan 14;12(2):598. doi: 10.3390/su12020598

